# Integrated Clinical-Genomic Reanalysis Identifies Diagnostic Gaps and Candidate Genes in a Pediatric Rare Disease Cohort

**DOI:** 10.64898/2026.09.24.26363679

**Authors:** Hyunjung Gu, Aniko Sabo, Sharayu V. Jangam, Hirokazu Hashimoto, Christina Y. Miyake, Bo Yuan, Charul Gijavanekar, Eric Venner, Viktoriya Korchina, Sarah H. Elsea, Donna M. Muzny, Elizabeth Mizerik, Blake Vuocolo, Poyyapakkam Srivaths, Lisa F. Satter, Sebastian Ochoa, Stephanie Sisley, Polle B. Mihindou, Oguz Kanca, Shinya Yamamoto, Zhandong Liu, Michael F. Wangler, Richard A. Gibbs, Hugo J. Bellen, Katherine Y. King

## Abstract

**Purpose:** Children with suspected rare genetic disorders often remain undiagnosed after exome, gene panel, or chromosomal microarray testing. We sought to define recurrent missed-diagnosis mechanisms and evaluate an integrated clinical-genomic reanalysis framework.

**Methods:** We studied 75 participants with rare or striking pediatric-onset phenotypes who remained undiagnosed after prior genetic testing. Trio genome sequencing evaluated SNVs, indels and copy-number variants. RNA sequencing was performed in 63 participants to assess aberrant expression and splicing, with targeted follow-up assays used selectively for prioritized variants. Clinician-genome scientist review integrated longitudinal phenotyping, physician input, and inheritance. Unresolved cases underwent research-level candidate prioritization.

**Results:** Clinically reportable findings were returned in 9, including two with multilocus findings: 5 explained the primary phenotype and 4, while not fully explanatory, were clinically relevant and informing surveillance, management, or family follow-up. Previously missed findings reflected technical limitations, including deep intronic splice-altering variants supported by transcriptome analysis and a genomic deletion, and interpretive limitations related to incomplete phenotype integration or narrowly scoped prior testing. Research-level analysis prioritized six candidate genes, including biallelic *ACHE* variants with reduced enzymatic activity.

**Conclusion:** Integrated clinical-genomic reanalysis framework identified diagnostic gaps, recovered care-informing findings, and prioritized candidate genes for rare disease discovery.

## INTRODUCTION

Rare genetic disease affects an estimated 3.5-5.9% of the population (1). The diagnostic odyssey for rare disease patients often takes years, contributing to suboptimal care, heavy healthcare utilization, and poor outcomes (2). For many families, genetic testing does not end the diagnostic odyssey; it becomes part of it. Exome sequencing (ES), targeted gene panels, chromosomal microarray analysis (CMA), and, increasingly, genome sequencing (GS) have improved molecular diagnosis for children with suspected rare genetic disorders (3–5). Yet access to testing, insurance coverage, and successful implementation of different testing modalities remain uneven across clinical settings (6, 7). As a result, many children remain without a definitive diagnosis despite extensive subspecialty evaluation and repeated genetic testing, creating a substantial and ongoing clinical burden (3, 5).

Among patients who remain undiagnosed, prior genetic testing histories often reflect more than a collection of negative reports. A nondiagnostic result may arise from gaps in what was measured, what was analyzed, or what could be interpreted at the time (5, 8). Some findings fall outside the technical scope of earlier assays, including noncoding, structural, or copy number variants (4, 5, 8). Others may be missed because analytic pipelines, variant-prioritization tools, prediction scores, or reporting thresholds differ across laboratories and evolve over time (4, 5, 9). Even when a variant is technically detectable, interpretation may remain limited by incomplete gene-disease knowledge, uncertain gene-phenotype correlations, narrow testing indications, or insufficient supporting evidence from longitudinal phenotyping, family history, segregation data and functional evidence (8–10). Thus, a prior negative or inconclusive report could later be resolved by different forms of reanalysis.

It can be difficult to determine the most informative next step for patients who remain undiagnosed after prior testing. In some cases, the unresolved question is primarily technical, requiring assessment of variant classes or molecular consequences not captured by prior assays; in others, the key barrier is interpretive, requiring updated phenotyping, inheritance assessment, transcript-level evidence, functional support, or multidisciplinary review (5, 8–11). A clinically useful reanalysis strategy therefore begins with structured review of prior testing and clinical context, followed by a multimodal molecular approach tailored to the specific unresolved question and coordinated interpretation to guide clinical return or research follow-up (8–12).

Here, we studied 75 participants referred by general pediatricians and pediatric subspecialists at a tertiary pediatric hospital for rare, striking, or otherwise unexplained pediatric-onset phenotypes; all lacked a confirmed molecular diagnosis at enrollment, including the majority who had undergone prior molecular or cytogenetic testing. We performed trio GS in all families, RNA sequencing (RNA-seq) in a subset, and targeted molecular follow-up for selected variants, combined with iterative clinician–genome scientist collaborative review incorporating longitudinal phenotyping and direct communication with treating physicians. Together, this approach integrated genomic, transcript-level, and clinical evidence to support clinical interpretation and candidate discovery. The goal was not only to measure additional diagnostic yield, but to define why prior testing remained nondiagnostic, identify which analytic and clinical-review steps contributed to interpretation, and show how unresolved cases can be guided toward clinical resolution or structured research discovery in a real-world setting.

## MATERIALS AND METHODS

The overall study workflow, including recruitment, clinical interpretation, return of results, and downstream research analysis, is summarized in Figure 1. The main methods are summarized below, with detailed descriptions of participant enrollment, sequencing protocols, bioinformatic pipelines, RNA-seq analysis, exon-trapping assays, and functional studies provided in the Supplementary Methods.

**Figure 1.**
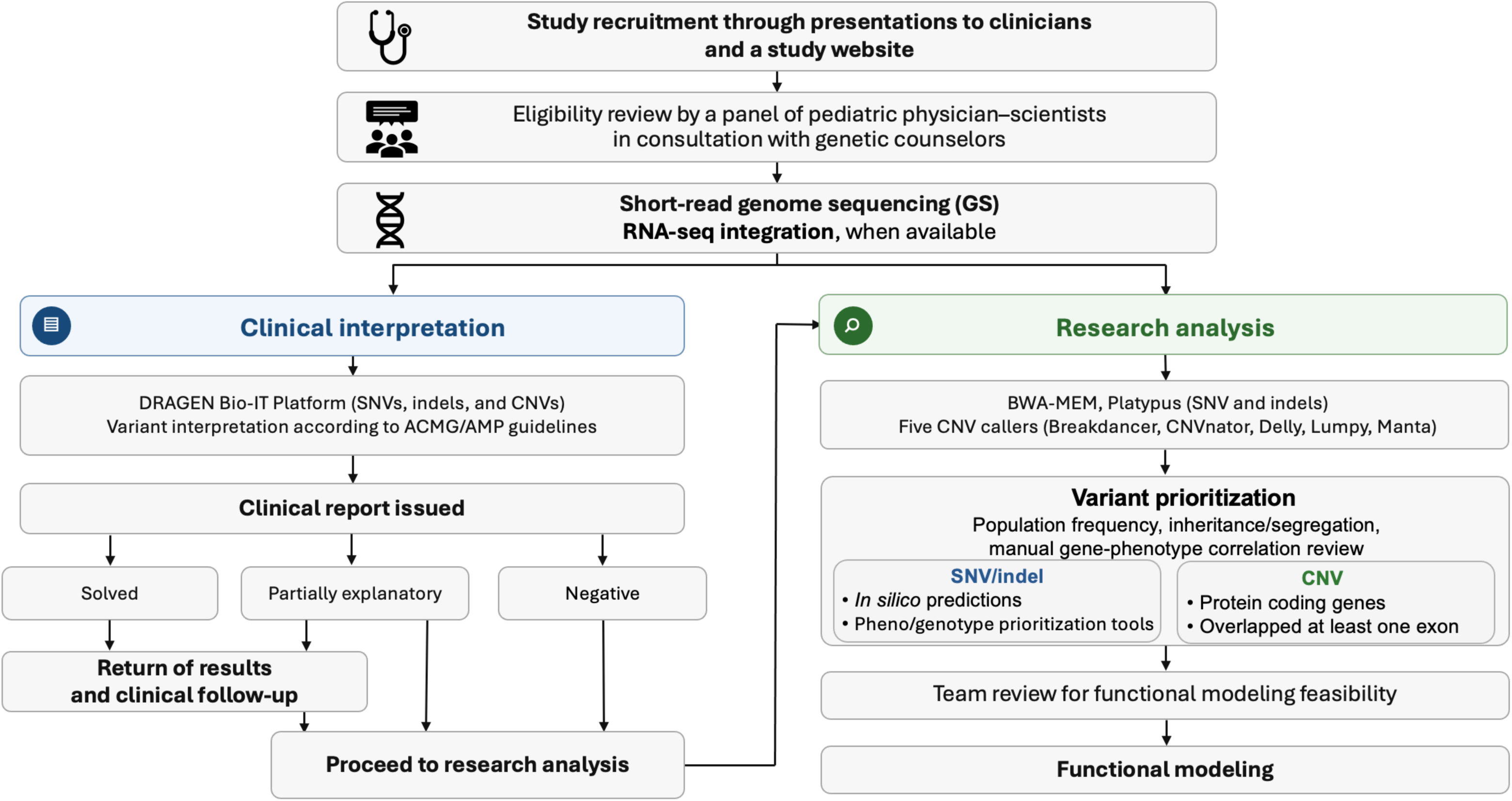
Study design and integrated clinical-genomic reanalysis workflow. Participants were recruited through presentations to clinicians at a tertiary pediatric hospital and a study website, followed by eligibility review by pediatric physician-scientists in consultation with genetic counselors. Enrolled participants underwent short-read GS, with RNA-seq integration when available. Clinical interpretation was performed using a clinical reporting pipeline, with variant interpretation according to the ACMG/AMP guidelines and return of clinically reportable findings. Cases with negative or not fully explanatory findings proceeded to research-level analysis, including SNV/indel and CNV prioritization, RNA-seq integration when available, team review for functional modeling feasibility, and functional follow-up of selected candidates. Abbreviations: ACMG/AMP, American College of Medical Genetics and Genomics/Association for Molecular Pathology; CMA, chromosomal microarray; CNV, copy number variants; ES, exome sequencing; GS, genome sequencing; RNA-seq, RNA sequencing; SNV, single nucleotide variant

### 1. Study population and clinical data collection

Participants were referred to the study by pediatricians and pediatric subspecialists at a tertiary pediatric hospital. The study was advertised through presentations to clinicians and through study information posted in clinical areas. Referrals were submitted through a HIPAA-compliant REDCap database and reviewed by a panel of pediatric physician-scientists in consultation with genetic counselors to determine study eligibility. Inclusion criteria included rare or striking disease presentations, multisystem involvement, and availability of biological parents for trio genetic testing. Exclusion criteria included a known or suspected genetic etiology explaining the disease presentation, prior enrollment in another study with an overlapping approach, or weak evidence for a genetically based syndrome. Patient enrollment and sample collection were conducted at the participating hospital. Written informed consent was obtained from all patients or their legal guardians. Clinical information was obtained from referral data, medical record review, Human Phenotype Ontology (HPO) terms provided by the study team, and, when needed, direct communication with treating physicians. Prior testing history was summarized using the first recorded genetic test available for each case. When prior testing was performed in an affected family member as part of the same diagnostic evaluation, the testing category was assigned at the case/family level. Intervals from the first recorded genetic test to study enrollment were compared between participants with and without newly identified clinically relevant findings using descriptive statistics and a two-sided Mann-Whitney U test.

### 2. Genome sequencing and analytic framework

Genome sequencing (GS) was performed for all enrolled participants and available family members. In principle, each family was analyzed as a trio, except for two duo families consisting of the participant and mother; affected siblings were included when available. Sequencing data were processed through complementary clinical and research pipelines, reflecting the dual goals of clinical reporting and research discovery. The clinical pipeline focused on identifying reportable variants in established disease-associated genes, whereas research-level analysis was used to prioritize candidate genes and variants in cases that remained negative or not fully explanatory after clinical interpretation.

### 3. RNA sequencing integration and targeted transcript-level follow-up

RNA sequencing (RNA-seq) was performed on peripheral blood from 63 participants to assess aberrant expression and splicing. RNA-seq served as the primary transcript-level approach and was used to evaluate expression and splicing outliers across the cohort and to support interpretation of selected candidate genes or variants prioritized from GS. Expression and splicing outliers were identified using DROP (v1.1.4) (13) with the OUTRIDER and FRASER modules, respectively, followed by visual review in Integrative Genomics Viewer (IGV) (14). Targeted exon-trapping assays were performed selectively for two prioritized deep intronic variants to assess predicted splice effects.

### 4. Clinical interpretation and return of results

For clinical reporting, raw GS reads were analyzed using the Illumina DRAGEN Bio-IT Platform (15), followed by calling of SNVs, indels, and copy-number variants. Variants were interpreted by the clinical team according to ACMG/AMP guidelines (16) in the context of the participant’s phenotype, inheritance pattern, and family history. Clinical reports were issued through the study clinical team. Findings considered solved or otherwise clinically relevant but not fully explanatory were returned to treating physicians and families, with clinical follow-up coordinated as appropriate. Cases with negative or not fully explanatory findings proceeded to research-level analysis.

### 5. Research-level analysis and candidate prioritization

Research-level analysis was performed for cases without a fully explanatory clinical diagnosis. SNVs/indels and CNVs were analyzed using complementary research pipelines, including BWA-MEM (17)/Platypus (18)-based SNV/indel analysis, multi-caller CNV assessment, variant annotation, *in silico* prediction, and AI-assisted prioritization. Candidate findings were reviewed based on rarity, inheritance or segregation, predicted deleteriousness, gene function, phenotype relevance, prior model-organism or human evidence, related biological mechanisms, and RNA-seq support when available. Final candidate genes were selected through collaborative review by the clinical and genomic analysis teams based on suitability for further investigation and feasibility for functional modeling. Consensus-prioritized candidate genes were submitted to GeneMatcher (19) and queried in available rare disease resources to identify additional individuals with overlapping phenotypes and candidate variants in the same genes.

### 6. Functional assessment of *ACHE* variants

*ACHE* variants prioritized through research analysis were evaluated using a cell-based acetylcholinesterase activity assay. Human *ACHE* reference and variant constructs were generated and transfected into HEK293T cells, and secreted acetylcholinesterase activity was measured in culture medium using a fluorescent activity assay. Experiments were performed in triplicate, with no-transfection and lacZ controls. Fluorescent activity measurements were compared using a two-tailed unpaired t test in GraphPad Prism.

## RESULTS

### 1. Cohort characteristics and prior testing history

We analyzed 75 participants with rare or striking disease presentations and no confirmed molecular diagnosis at enrollment (Figure 1, Table 1). The median age at enrollment was 12 years (age categories: 0–4 to 25–29 years), and the cohort included 35 males (46.7%) and 40 females (53.3%). Based on self-reported race and ethnicity, 32 participants (42.7%) identified as Hispanic/Latino and 43 (57.3%) as non-Hispanic/Latino. These characteristics reflect the diverse referral population of the participating pediatric hospital. Most participants exhibited multisystem involvement (61.3%), with frequent cardiovascular, hematologic/immunologic, neurologic/psychiatric, gastrointestinal, and musculoskeletal manifestations.

**Table 1.** Characteristics of the study population (n=75)

| **Characteristics** |  | **Number of probands (%)** |
| --- | --- | --- |
| Age at the time of enrollment | **Median age: 12 years** (age categories: 0–4 to 25–29 years) | **75 (100%)** |
| Sex | Male | 35 (46.7%) |
|  | Female | 40 (53.3%) |
| Race/Ethnicity | **Non-Hispanic/Latino** | 43 (57.3%) |
|  | Asian | 7 (9.3%) |
|  | Black | 2 (2.7%) |
|  | White | 34 (79.1%) |
|  | Other | 0 (0.0%) |
|  | **Hispanic/Latino** | 32 (42.7%) |
|  | White | 32 (100.0%) |
| Referred Subject Phenotypes | **Multisystem Involvement** | 46 (61.3%) |
|  | Cardiovascular | 35 (46.7%) |
|  | Dermatologic | 11 (14.7%) |
|  | ENT | 8 (10.7%) |
|  | Endocrinologic | 10 (13.3%) |
|  | Gastrologic | 22 (29.3%) |
|  | Gynecologic | 6 (8.0%) |
|  | Hematologic/Immunologic | 24 (32.0%) |
|  | Musculoskeletal | 19 (25.3%) |
|  | Neurologic/Psychiatric | 22 (29.3%) |
|  | Ophthalmologic | 7 (9.3%) |
|  | Pulmonary/Respiratory | 15 (20.0%) |
|  | Urologic/Renal | 6 (8.0%) |
|  | Other | 6 (8.0%) |
| Previous Genetic Testing | **YES** | 59 (78.7%) |
|  | Genome Sequencing | 6 (8.0%) |
|  | Exome Sequencing | 36 (48.0%) |
|  | Multiple Gene Panel Sequencing | 31 (41.3%) |
|  | Chromosomal Microarray Analysis (CMA) | 26 (34.7%) |
|  | Other | 18 (24.0%) |
|  | **NO** | 16 (21.3%) |

Prior clinical genetic testing was common and heterogeneous (Table 1). Overall, 59 participants (78.7%) had undergone at least one prior clinical genetic test, including genome sequencing (8.0%), exome sequencing (48.0%), multi-gene panel testing (41.3%), chromosomal microarray analysis (34.7%), or other genetic, biochemical or cytogenetic testing (24.0%). The remaining 16 participants (21.3%) had no documented prior genetic testing. Prior evaluations frequently involved overlapping modalities; 25 participants (33.3%) had undergone ES and/or GS in combination with CMA, often as part of broader diagnostic workups that included additional targeted or adjunct testing. Among 59 participants with prior testing, dates of the first recorded genetic test were available for 56 participants. The median time from the first genetic test to study enrollment was 879 days (interquartile range (IQR), 412-2346 days; range, 55-6536 days), underscoring the prolonged diagnostic odyssey in this cohort. When stratified by reanalysis outcome, participants (n=9) with newly identified clinically relevant findings had a longer median interval than those without positive findings (n=47) (1566 [positive] vs. 874 [negative] days; IQRs, 514-2207 vs. 374-2346 days; ranges, 55-3716 vs. 105-6536 days), although this difference was not statistically significant and the IQRs overlapped (Supplementary Figure 1). Initial testing approaches were heterogeneous, most commonly consisting of gene panels or CMA, whereas ES or GS accounted for a smaller proportion of first recorded tests. Notably, none of the participants with clinically relevant findings had ES or GS as their first recorded test.

### 2. Clinical analysis identified reportable findings in 9 participants

Clinical analysis identified newly reportable findings in 9 of 75 participants (12.0%) (Table 2). Detailed variant information for these participants is provided in Supplementary Table 1, and additional clinical summaries for individual cases are provided in the Supplementary File.

**Table 2.**
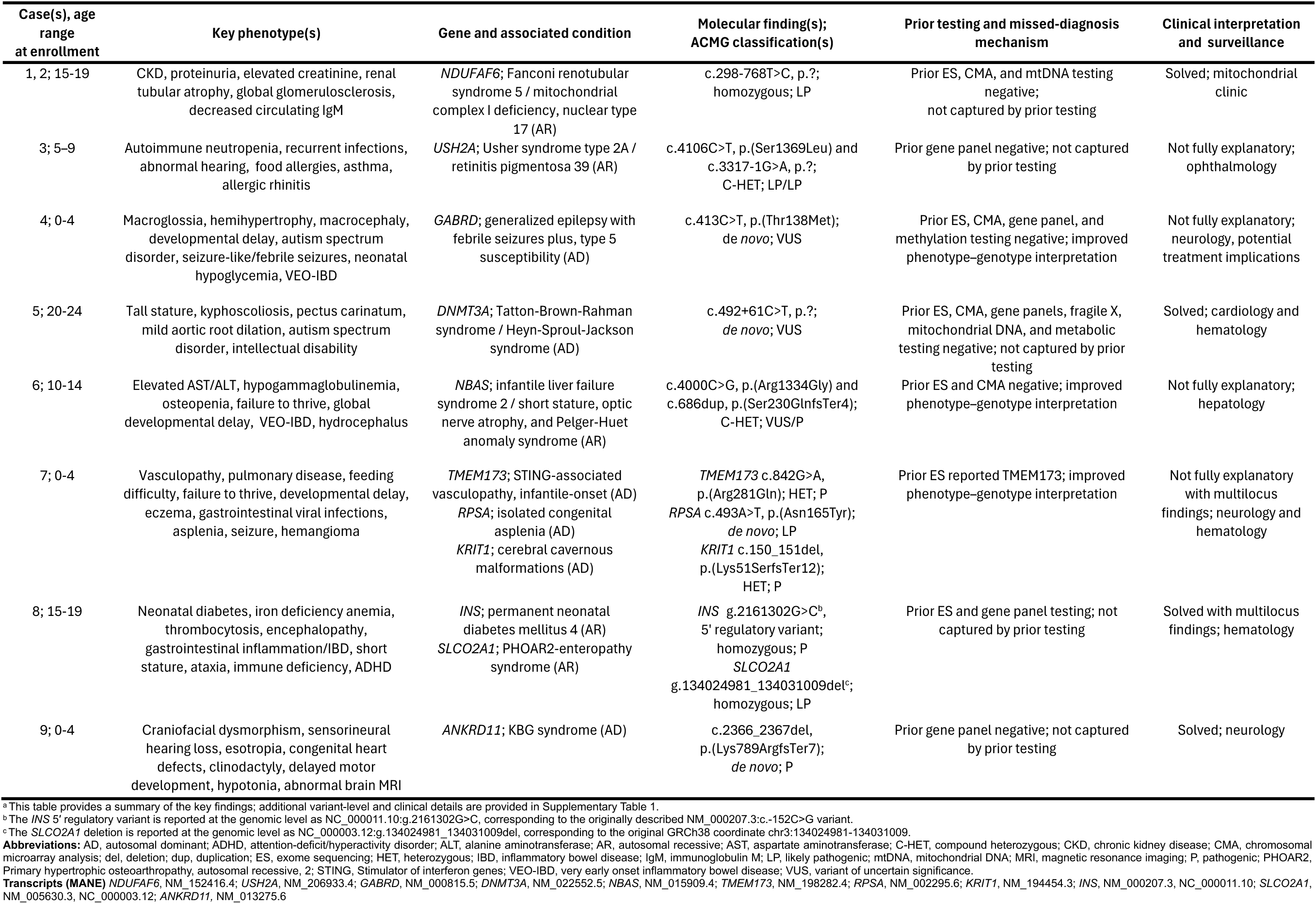
Newly diagnosed cases (n=9) ^a^.

All 9 participants had previously undergone genetic testing, including ES or GS in 7 participants (Figure 2). Seven had no diagnostic findings reported in prior testing. The remaining 2 participants had pathogenic or likely pathogenic variants identified previously; however, those findings explained only a subset of the patients’ phenotypes, and the cases were referred for further analysis to identify additional genetic findings that could account for the primary clinical features.

**Figure 2.**
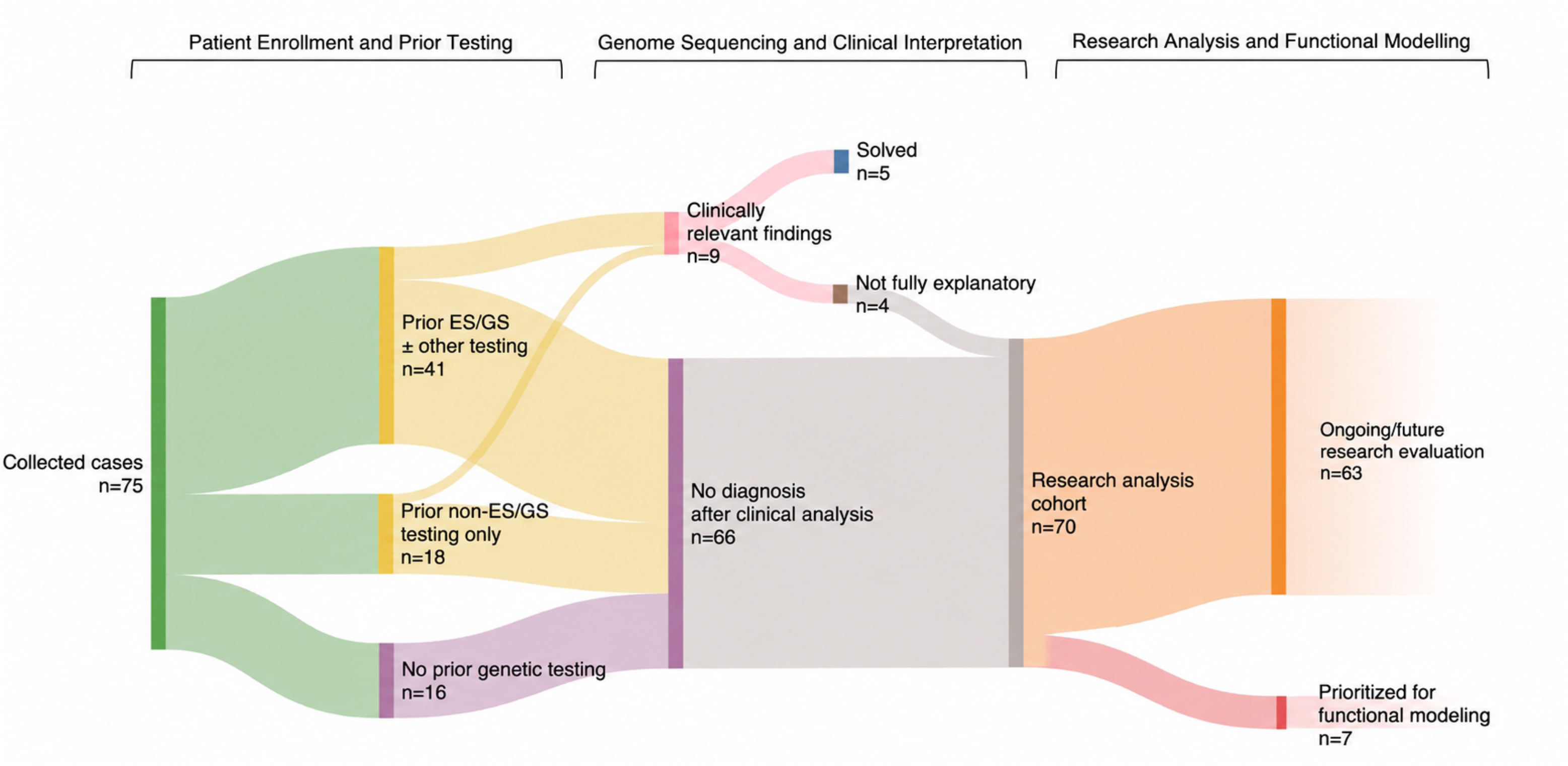
Cohort-level flow from enrollment and prior testing to clinical and research outcomes. The diagram shows the distribution of 75 enrolled participants according to prior testing history, clinical interpretation, clinically reportable findings, and downstream research analysis. At enrollment, 41 unique participants had prior ES or GS with or without other testing, 18 had prior panel, CMA, or other testing only, and 16 had no documented prior genetic testing. One participant had undergone both ES and GS. Clinical interpretation identified clinically reportable findings in 9 participants, including 5 findings that explained the primary phenotype and 4 clinically relevant findings that were not fully explanatory. Cases without a fully explanatory clinical diagnosis proceeded to the research analysis cohort, where selected cases were prioritized for functional modeling or ongoing/future research evaluation. Abbreviations: CMA, chromosomal microarray; ES, exome sequencing; GS, genome sequencing

Among the 9 participants with clinically reportable findings, 5 had findings were considered fully explanatory for the primary phenotype. The remaining 4 had findings that were clinically relevant but not fully explanatory for the referral phenotype (Figure 2). These included findings that accounted for a subset of clinical features or identified medically relevant risks requiring surveillance or follow-up but did not explain the full clinical spectrum or primary referral indication.

### 3. Missed findings reflected technical and interpretive limitations of prior testing

Non-reporting in prior testing occurred for two primary reasons. In 6 cases, the variant was not captured or detectable by the testing methodology used at the time, such as deep intronic variants, as well as a large genomic deletion missed by prior exome-based testing. In the remaining 3 cases, the variant was technically detectable but was not interpreted as diagnostic, reflecting incomplete integration of the clinical phenotype, family history, or the broader clinical question at the time of prior testing. The newly detected variants included coding or canonical splice-site variants, deep intronic splicing variants, and a large genomic deletion (Table 2, Supplementary Table 1).

Representative cases illustrate how different data layers contributed to interpretation. In two siblings with progressive renal insufficiency (Cases 1 and 2), prior ES, CMA, and mitochondrial DNA testing were nondiagnostic. GS identified a homozygous deep intronic *NDUFAF6* (HGNC:28625) c.298-768T>C variant (20). RNA-seq from both siblings showed near-complete loss of the normal exon 2-3 junction compared with parental samples and additional controls, supporting aberrant splicing of a major *NDUFAF6* transcript.

In a patient with overgrowth, aortic root dilation, connective tissue features, and intellectual disability (Case 5), GS identified a *de novo* deep intronic *DNMT3A* (HGNC:2978) variant with a strong SpliceAI donor-gain prediction (0.98). Although patient-derived RNA-seq was not available for this case, targeted exon trapping enabled functional assessment for the predicted splice effect. The assay demonstrated creation of a novel splice donor site that extended exon 5 from 44 bp to 103 bp, consistent with the *in-silico* prediction. The aberrant transcript is predicted to introduce p.Lys166Serfs*79 and likely undergo nonsense-mediated decay.

In another patient with refractory inflammatory bowel disease, hypertrophic osteoarthropathy, and myelofibrosis (Case 8), GS-based CNV analysis identified a large *SLCO2A1* (HGNC:10955) deletion that had not been detected by prior ES or inflammatory bowel disease panel testing.

Other findings reflected interpretive rather than purely technical advances. In these cases, the relevant variants were likely within the analytic scope of prior testing but were not recognized as clinically meaningful because earlier analyses were directed toward a narrower clinical presentation or lacked the longitudinal phenotypic context available during reanalysis. For example, in the participant with *NBAS* (HGNC:15625)-associated disease (Case 6), liver enzyme abnormalities and longitudinal hepatic features were not incorporated into the prior interpretation, whereas their inclusion during reanalysis supported the relevance of the *NBAS* finding and prompted hepatology follow-up. In another participant with biallelic *USH2A* (HGNC:12601) variants (Case 3), the finding did not explain the primary referral indication but identified a clinically relevant risk requiring ophthalmologic surveillance. Together, these examples illustrate how iterative review involving both the clinical and genomic analysis teams allowed broader phenotypic information to be incorporated, earlier assumptions to be revisited, and new genotype-phenotype correlations to be identified.

### 4. Returned findings informed clinical management and surveillance

Return of results translated into changes in clinical care in nine participants. Newly established diagnoses prompted referrals to relevant subspecialties, including mitochondrial clinic, neurology, pulmonary, cardiology, hematology, hepatology, and ophthalmology, and led to surveillance strategies tailored to the underlying molecular diagnosis. Examples included cardiac and hematologic surveillance for Tatton-Brown-Rahman syndrome in the participant with a *de novo DNMT3A* deep intronic variant (Case 5), neurologic monitoring and seizure surveillance for KBG syndrome in the participant with a *de novo ANKRD11* (HGNC:21316) loss-of-function variant (Case 9), hepatology follow-up in the participant with *NBAS*-associated disease (Case 6), and ophthalmologic surveillance in the participant with biallelic *USH2A* variants despite the absence of current visual symptoms (Case 3).

Returned findings also informed management beyond surveillance. Two multilocus cases illustrate this point. In Case 7, reanalysis in the participant with an existing *TMEM173* finding (HGNC:27962), additionally identified findings involving *RPSA* (HGNC:6502) and *KRIT1* (HGNC:1573), which informed family genetic screening and imaging follow-up for *KRIT1*-related cerebral cavernous malformations. In Case 8, the participant had a prior *INS* finding (HGNC:6081), and reanalysis identified a large *SLCO2A1* deletion (HGNC:10955). The genetic diagnosis prompted ankle radiography confirming hypertrophic osteoarthropathy and bone marrow biopsy confirming WHO grade 1 myelofibrosis.

Together, these outcomes show that returned findings affected care not only by establishing molecular diagnoses, but also by guiding downstream evaluation, clarifying complex phenotype attribution, and refining risk assessment for patients and relatives.

### 5. Research-level prioritization identified candidates for functional follow-up

Among cases that remained without a fully explanatory clinical diagnosis (n=70), research-level analyses prioritized 7 cases involving 6 candidate genes for further investigation (Figure 2). These candidates are undergoing functional follow-up studies to assess biological plausibility and potential disease mechanisms.

As a representative example, we identified compound heterozygous variants in *ACHE* (HGNC:108), which encodes acetylcholinesterase, a well-characterized enzyme that terminates cholinergic neurotransmission and carries the Yt blood group antigen on erythrocytes (21, 22). *ACHE* was considered a biologically plausible candidate because it encodes a highly evolutionarily conserved enzyme critical for cholinergic neurotransmission (23), and acetylcholinesterase disruption causes severe neuromuscular or neurologic phenotypes in model organisms (24–27), although *ACHE* has not been established as a Mendelian disease gene in humans.

The participant had global developmental delay, refractory epilepsy, autism, congenital nystagmus, hypotonia, scoliosis, prolonged QTc interval, and autonomic or cholinergic-like features, including pinpoint pupils, drooling, and abnormal anesthesia reactions. The phenotype partially overlapped with cholinergic excess (28) and congenital myasthenic syndrome phenotypes (29) but showed a distinct chronic multisystem pattern (Supplementary Table 2). Research analysis identified two variants inherited in trans: a frameshift variant (NM_000665.5:c.11dup, p.(Gln5AlafsTer28)), predicted to result in loss of function, and a missense variant (NM_000665.5:c.499C>G, p.(Arg167Gly)) with a CADD score of 26.7 (REVEL 0.523, AlphaMissense ambiguous). Within the family, genetic material from a similarly affected individual who died suddenly during sleep during adolescence was unavailable for segregation analysis; an individual carrying only the missense variant was unaffected.

Functional assessment in HEK293T cells demonstrated markedly reduced acetylcholinesterase activity for both *ACHE* variants compared with cells expressing reference ACHE, with the missense variant retaining slightly higher residual activity than the frameshift allele (Figure 3). These findings support a hypomorphic model in which a null allele in combination with a minimally functional missense allele results in severely reduced, but not absent, acetylcholinesterase activity (Figure 3). The absence of homozygous predicted loss-of-function *ACHE* variants in population databases is consistent with intolerance to complete loss of ACHE function (pLI 0.81, LOEUF 0.37). However, because no additional matched participants have been identified through GeneMatcher or other rare-disease matching resources to date, functional validation in additional affected individuals has not yet been completed to establish causality.

**Figure 3.**
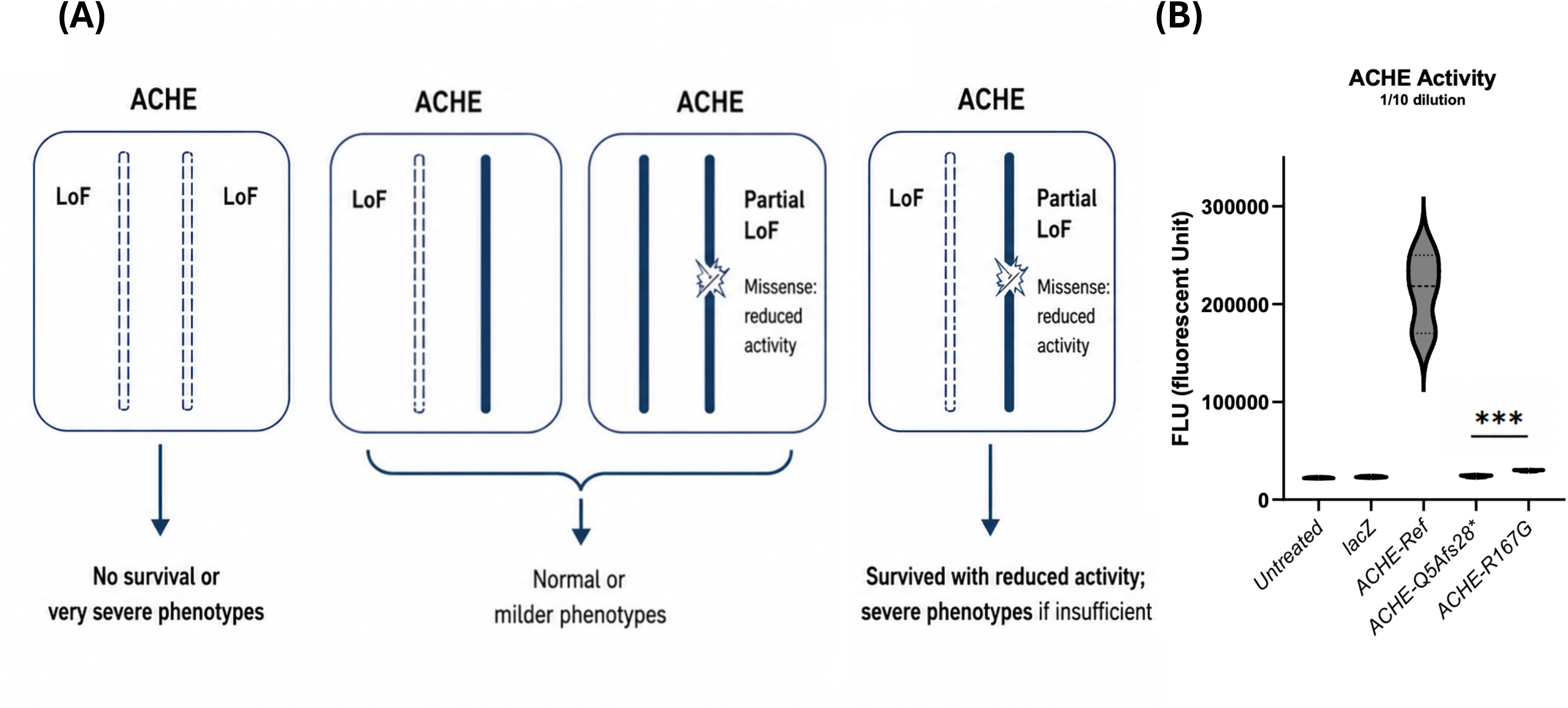
Predicted genotype-phenotype relationship and functional assessment of *ACHE* variants. Left, schematic model illustrating the proposed genotype–phenotype relationship for *ACHE* variants. Biallelic complete loss-of-function (LoF/LoF) is predicted to be incompatible with survival or associated with very severe phenotypes, whereas heterozygous carriers with one functional allele (LoF/WT or WT/partial LoF) are expected to have normal or milder phenotypes. In contrast, compound heterozygosity for a null allele and a hypomorphic missense allele (LoF/partial LoF) may permit minimal residual ACHE activity, allowing survival but resulting in severe multisystem manifestations, as observed in the proband. Right, functional assessment of ACHE enzymatic activity in HEK293T cells transfected with reference or variant ACHE constructs. Secreted ACHE activity was quantified from culture supernatants using a fluorescence-based enzymatic assay. Compared with the reference construct (ACHE-Ref), both patient-associated variants (ACHE-Q5Afs27* and ACHE-Arg167Gly) showed markedly reduced enzymatic activity, with the missense variant retaining slightly higher residual activity than the frameshift/null variant. These findings support a loss-of-function mechanism and suggest that minimal residual ACHE activity may underlie survival in the setting of severe cholinergic phenotypes. \*\**p* < 0.001; FLU, fluorescence units; LoF, loss of function; WT, wild type.

## DISCUSSION

In this study, we evaluated 75 participants with rare or striking pediatric-onset phenotypes who lacked a confirmed molecular diagnosis at enrollment, despite extensive clinical evaluation and, in many cases, prior genetic testing. Integrated clinical-genomic reanalysis identified clinically reportable findings in 9 participants, including 5 findings that explained the primary phenotype and 4 clinically relevant findings that were not fully explanatory but informed surveillance, management, or follow-up.

Prior experience from the Undiagnosed Diseases Network (UDN) and other rare disease reanalysis efforts has shown that unresolved cases often benefit from multimodal evaluation and periodic reassessment as clinical and molecular evidence evolves (30, 31). The cohort adds a complementary perspective by focusing on pediatric participants who had already passed through heterogeneous real-world clinical testing pathways yet remained without a confirmed molecular diagnosis or with an incompletely explained phenotype. In this setting, reanalysis clarified the residual gaps left by prior testing, including technical blind spots of earlier assays, incomplete phenotype integration, and findings whose clinical value depended on management implications rather than full explanation of the presenting phenotype. This perspective frames reanalysis as a process of recontextualizing prior results within updated clinical and molecular evidence, rather than simply escalating to a broader assay.

The interval analysis adds a temporal dimension to this interpretation. In the cohort, participants with clinically relevant findings had a longer median interval from first recorded genetic testing to reanalysis than those without such findings, with median intervals of approximately 4 years and 2 years, respectively. Although this difference did not reach statistical significance, its direction is clinically plausible: these participants had also entered the diagnostic trajectory through more limited initial testing, and over an interval of several years, both the methods available for genetic testing and the interpretive frameworks applied to their results can advance considerably. As a result, an evaluation that was once unrevealing may become clinically informative on reassessment. In this context, while based on a limited cohort, the data support using 4 years’ elapsed time as a reasonable interval for reanalysis of unresolved cases. We expect such reassessment to be particularly valuable for individuals whose prior testing was limited in scope, predates current analytic approaches, or no longer adequately addresses the evolving clinical presentation.

Returned findings carried clinical value in different ways. Some findings unified the primary clinical presentation and provided diagnostic closure. Others were clinically relevant in cases where the phenotype was only partially explained by the molecular finding or could have been interpreted as phenotypic expansion. In addition, some participants had multilocus results in which reanalysis identified clinically relevant findings beyond a prior molecular result. Together, these examples highlight that the clinical value of reanalysis lies not only in identifying a molecular diagnosis, but also in clarifying phenotype attribution in complex presentations.

Prior studies of multilocus variation and blended phenotypes have shown that diagnostic evaluation is not necessarily complete after an initial molecular finding (32, 33). Consistent with this concept, this cohort included examples in which reanalysis helped distinguish findings that explained the primary phenotype from additional clinically relevant findings that informed surveillance, counseling, or follow-up. Because clinical expertise is often needed to untangle the relationship between phenotypes observed in the clinic and molecular diagnoses identified by genome-wide assays (33), this perspective supports clinician-scientist review to determine which clinical features are attributable to which molecular findings or other clinical factors, particularly when a result is clinically relevant but might otherwise be overlooked.

The same framework also created a pathway for research-prioritized candidates in cases that remained unresolved after clinical interpretation. This is an important output of pediatric rare disease reanalysis. Cases without sufficient evidence for clinical return can still generate testable gene-disease hypotheses when variant-level evidence, phenotype relevance, segregation, molecular data, and functional feasibility converge. The family with *ACHE* variants illustrates this role. Although *ACHE* has not been established as a Mendelian disease gene in humans, the combination of a highly suggestive phenotype, biallelic variants, segregation data from available family members, population-level constraint, and markedly reduced acetylcholinesterase activity nominates *ACHE* as a biologically plausible candidate. Additional affected individuals and in vivo functional validation will be needed to establish causality, but this example illustrates how clinically anchored research analysis can generate testable hypotheses and contribute to the broader effort to define the genetic basis of rare disease.

Several limitations should be considered. Although this cohort was deeply phenotyped and uniformly evaluated with trio genome sequencing, certain variant classes may remain incompletely assessed, including low-level mosaicism, mitochondrial DNA variants, repeat expansions, complex rearrangements, regulatory variants, and structural variants beyond the CNVs evaluated here. In addition, research-prioritized candidates will require additional affected individuals and functional validation to establish definitive gene-disease relationships.

This study represents the analysis of a highly challenging cohort seeking molecular diagnosis, enriched for previously tested and clinically complex cases. These unresolved pediatric rare disease cases represent a heterogeneous group, with prior nondiagnostic evaluations reflecting different combinations of technical, interpretive, and clinical-context limitations. We showed that structured clinical-genomic reanalysis can identify clinically reportable findings, clarify mechanisms underlying prior nondiagnostic results, and prioritize candidates for research follow-up. These findings support a practical framework to guide both patient care and candidate discovery in children with suspected genetic disorders who remain without a confirmed molecular diagnosis.

## Data Availability statement

All data are included within the paper or its supplemental information. Raw genome sequencing and raw RNA sequencing data cannot be shared given the consent under which the affected individuals were recruited. Variants of clinical relevance in this manuscript have been deposited in ClinVar under accession numbers SCV007538406-SCV007538420.

## Acknowledgement

We sincerely thank the patients and their families for their participation in this research.

## Author Contributions

Conceptualization: R.A.G., H.J.B., K.Y.K.

Data curation: K.Y.K., M.F.W., A.S., H.G.

Formal analysis: H.G., A.S., B.Y., C.G., S.H.E.

Funding acquisition: R.A.G., H.J.B., K.Y.K.

Investigation: C.Y.M., K.Y.K., M.F.W., P.S., L.F.S., S.O.G., S.S., P.B.M., E.M., B.V., S.V.J., H.H., O.K., S.Y.

Methodology: V.K., D.M., A.S., H.G., E.V.,

Project administration: P.B.M.

Software: A.S., H.G., E.V., Z.L.

Validation: M.F.W., S.V.J., H.H., O.K., S.Y.

Visualization: H.G., S.V.J., H.H.

Writing-original draft: H.G., A.S., R.A.G., K.Y.K., M.F.W.

Writing-review & editing: All authors.

## Ethics declaration

This study was approved by the Institutional Review Board of Baylor College of Medicine (H-53886). Participants were referred by healthcare professionals through a Health Insurance Portability and Accountability Act (HIPAA)-compliant REDCap database and enrolled at Texas Children’s Hospital. Written informed consent was obtained from all participants or their legal guardians prior to inclusion in the study.

## Supplementary Information

**Supplementary File** Detailed methods and supplementary references

**Supplementary Table 1** Detailed variant information for participants with clinically reportable findings (n=9).

**Supplementary Table 2** Comparison of clinical features in the case in this study and representative acetylcholinesterase-related or cholinergic disorders

**Supplementary Figure 1** Interval to reanalysis by outcome group

## References

1. Nguengang Wakap S, Lambert DM, Olry A, Rodwell C, Gueydan C, Lanneau V, et al. Estimating cumulative point prevalence of rare diseases: analysis of the Orphanet database. Eur J Hum Genet. 2020;28(2):165–73.

2. Glaubitz R, Heinrich L, Tesch F, Seifert M, Reber KC, Marschall U, et al. The cost of the diagnostic odyssey of patients with suspected rare diseases. Orphanet J Rare Dis. 2025;20(1):222.

3. Marwaha S, Knowles JW, Ashley EA. A guide for the diagnosis of rare and undiagnosed disease: beyond the exome. Genome Med. 2022;14(1):23.

4. Wojcik MH, Lemire G, Berger E, Zaki MS, Wissmann M, Win W, et al. Genome Sequencing for Diagnosing Rare Diseases. N Engl J Med. 2024;390(21):1985–97.

5. Wojcik MH, Reuter CM, Marwaha S, Mahmoud M, Duyzend MH, Barseghyan H, et al. Beyond the exome: What’s next in diagnostic testing for Mendelian conditions. Am J Hum Genet. 2023;110(8):1229–48.

6. Zion TN, Berrios CD, Cohen ASA, Bartik L, Cross LA, Engleman KL, et al. Insurance denials and diagnostic rates in a pediatric genomic research cohort. Genet Med. 2023;25(5):100020.

7. Robillard JM, Feng TL, Kabacinska K. Access to genetic testing for rare diseases: Existing gaps in public-facing information. World Med Health Policy. 2021;13(3):518–25.

8. Ewans LJ, Schofield D, Shrestha R, Zhu Y, Gayevskiy V, Ying K, et al. Whole-exome sequencing reanalysis at 12 months boosts diagnosis and is cost-effective when applied early in Mendelian disorders. Genet Med. 2018;20(12):1564–74.

9. Bullich G, Matalonga L, Pujadas M, Papakonstantinou A, Piscia D, Tonda R, et al. Systematic Collaborative Reanalysis of Genomic Data Improves Diagnostic Yield in Neurologic Rare Diseases. J Mol Diagn. 2022;24(5):529–42.

10. Matalonga L, Hernandez-Ferrer C, Piscia D, Solve RDSNViwg, Schule R, Synofzik M, et al. Solving patients with rare diseases through programmatic reanalysis of genome-phenome data. Eur J Hum Genet. 2021;29(9):1337–47.

11. Yepez VA, Gusic M, Kopajtich R, Mertes C, Smith NH, Alston CL, et al. Clinical implementation of RNA sequencing for Mendelian disease diagnostics. Genome Med. 2022;14(1):38.

12. Kremer LS, Bader DM, Mertes C, Kopajtich R, Pichler G, Iuso A, et al. Genetic diagnosis of Mendelian disorders via RNA sequencing. Nat Commun. 2017;8:15824.

13. Yépez VA, Mertes C, Mueller MF, Andrade DS, Wachutka L, Frésard L, et al. Detection of aberrant gene expression events in RNA sequencing data. 2019.

14. Robinson JT, Thorvaldsdottir H, Winckler W, Guttman M, Lander ES, Getz G, et al. Integrative genomics viewer. Nat Biotechnol. 2011;29(1):24–6.

15. Behera S, Catreux S, Rossi M, Truong S, Huang Z, Ruehle M, et al. Comprehensive genome analysis and variant detection at scale using DRAGEN. Nat Biotechnol. 2025;43(7):1177–91.

16. Richards S, Aziz N, Bale S, Bick D, Das S, Gastier-Foster J, et al. Standards and guidelines for the interpretation of sequence variants: a joint consensus recommendation of the American College of Medical Genetics and Genomics and the Association for Molecular Pathology. Genet Med. 2015;17(5):405–24.

17. Li H, Durbin R. Fast and accurate short read alignment with Burrows-Wheeler transform. Bioinformatics. 2009;25(14):1754–60.

18. Rimmer A, Phan H, Mathieson I, Iqbal Z, Twigg SRF, Consortium WGS, et al. Integrating mapping-, assembly- and haplotype-based approaches for calling variants in clinical sequencing applications. Nat Genet. 2014;46(8):912–8.

19. Sobreira N, Schiettecatte F, Valle D, Hamosh A. GeneMatcher: a matching tool for connecting investigators with an interest in the same gene. Hum Mutat. 2015;36(10):928–30.

20. Hartmannova H, Piherova L, Tauchmannova K, Kidd K, Acott PD, Crocker JF, et al. Acadian variant of Fanconi syndrome is caused by mitochondrial respiratory chain complex I deficiency due to a non-coding mutation in complex I assembly factor NDUFAF6. Hum Mol Genet. 2016;25(18):4062–79.

21. Getman DK, Eubanks JH, Camp S, Evans GA, Taylor P. The human gene encoding acetylcholinesterase is located on the long arm of chromosome 7. Am J Hum Genet. 1992;51(1):170–7.

22. Spring FA, Gardner B, Anstee DJ. Evidence that the antigens of the Yt blood group system are located on human erythrocyte acetylcholinesterase. Blood. 1992;80(8):2136–41.

23. Pickett MA, Dush MK, Nascone-Yoder NM. Acetylcholinesterase plays a non-neuronal, non-esterase role in organogenesis. Development. 2017;144(15):2764–70.

24. Hall JC, Kankel DR. Genetics of acetylcholinesterase in Drosophila melanogaster. Genetics. 1976;83(3 PT.2):517-35.

25. Behra M, Cousin X, Bertrand C, Vonesch JL, Biellmann D, Chatonnet A, et al. Acetylcholinesterase is required for neuronal and muscular development in the zebrafish embryo. Nat Neurosci. 2002;5(2):111–8.

26. Greenspan RJ, Finn JA, Jr., Hall JC. Acetylcholinesterase mutants in Drosophila and their effects on the structure and function of the central nervous system. J Comp Neurol. 1980;189(4):741–74.

27. Li B, Duysen EG, Volpicelli-Daley LA, Levey AI, Lockridge O. Regulation of muscarinic acetylcholine receptor function in acetylcholinesterase knockout mice. Pharmacol Biochem Behav. 2003;74(4):977–86.

28. Adeyinka A, Patel A, Kondamudi NP. Cholinergic Crisis. StatPearls. Treasure Island (FL)2026.

29. Ohno K, Brengman J, Tsujino A, Engel AG. Human endplate acetylcholinesterase deficiency caused by mutations in the collagen-like tail subunit (ColQ) of the asymmetric enzyme. Proc Natl Acad Sci U S A. 1998;95(16):9654–9.

30. Murdock DR, Rosenfeld JA, Lee B. What Has the Undiagnosed Diseases Network Taught Us About the Clinical Applications of Genomic Testing? Annu Rev Med. 2022;73:575–85.

31. Furuta Y, Tinker RJ, Hamid R, Cogan JD, Ezell KM, Oglesbee D, et al. A review of multiple diagnostic approaches in the undiagnosed diseases network to identify inherited metabolic diseases. Orphanet J Rare Dis. 2024;19(1):427.

32. Posey JE, Harel T, Liu P, Rosenfeld JA, James RA, Coban Akdemir ZH, et al. Resolution of Disease Phenotypes Resulting from Multilocus Genomic Variation. N Engl J Med. 2017;376(1):21–31.

33. Karaca E, Posey JE, Coban Akdemir Z, Pehlivan D, Harel T, Jhangiani SN, et al. Phenotypic expansion illuminates multilocus pathogenic variation. Genet Med. 2018;20(12):1528–37.

