## Supplementary File for "Integrated Clinical-Genomic Reanalysis Identifies Diagnostic Gaps and Candidate Genes in a Pediatric Rare Disease Cohort"

**(Supplementary Materials)**

**Supplementary Materials and Methods**

The overall study workflow, including recruitment, clinical interpretation, return of results, and downstream research analysis, is summarized in Figure 1.

1. **Study Population, Recruitment, and Outreach**

Individuals were referred to the study by pediatricians and pediatric subspecialists at a tertiary pediatric hospital. The study was advertised through direct presentations to clinicians and through study information and website links posted in clinical areas at the hospital in English, Spanish, and Vietnamese.

2. **Participant enrollment and sampling process**

The study was approved by the Institutional Review Board of the participating institution. Patients were referred by healthcare professionals through a Health Insurance Portability and Accountability Act (HIPAA)-compliant REDCap database. Referrals were reviewed by a panel of five pediatric physician-scientists in consultation with two genetic counselors to determine eligibility for the study. Inclusion criteria included rare or striking disease presentations, multisystem involvement, and availability of biological parents for trio genetic testing. Exclusion criteria included known or suspected genetic etiology of disease presentation, prior enrollment in another study with an overlapping approach (e.g. Undiagnosed Disease Network), or weak evidence for a genetically based syndrome (e.g. the phenotype is common and without additional family members with similar phenotypes).

Patient enrollment and sample collection were conducted at the participating hospital. Written informed consent was obtained from all patients and parents prior to participation. Trained medical translators were utilized as appropriate. The clinical team assembled Human Phenotype Ontology (HPO) terms for referred patients based on the medical record. Prior testing history was summarized using the first recorded genetic test available for each case. When prior testing was performed in an affected family member as part of the same diagnostic evaluation, the testing category was assigned at the case/family level. Intervals from the first recorded genetic test to study enrollment were compared between participants with and without newly identified clinically relevant findings using descriptive statistics and a two-sided Mann–Whitney U test. Box plots were generated in Python 3.10 with Matplotlib 3.10 and SciPy 1.15.

Blood samples for DNA and RNA sequencing were collected in PAXgene® tubes and transferred to the sequencing center at the participating institution, where they were processed according to standard protocols.

3. **Genomic sequencing pipelines**

1) Genome sequencing (GS)

Genome sequencing (GS) was performed to enable comprehensive detection of both coding and noncoding variants. Briefly, Genomic DNA was robotically purified from peripheral blood collected in PAXgene® Blood DNA tubes using the Revvity Chemagic™ Prime 8 automated platform. The purified DNA was fragmented by sonication, double-size selected with AmPure XP beads, and ligated to Illumina Dual-index barcode adapters. The adapter-ligated DNA was then purified with AmPure XP beads. Concentration of the final library was measured with the Kapa Library Quant kit.

Libraries were normalized and pooled for massively parallel sequencing. The library pools were subjected to sequence analysis on an Illumina NovaSeq 6000 platform for 150 bp paired end reads. The following quality control metrics of the sequencing data were achieved: QC1 Mean Coverage (Dragen) ≥38X with 97% of bases attaining at least 20X coverage; Q30 Bases (Dragen) ≥8E10; Mapped reads ≥99%; Estimated Contamination (Dragen) ≤1%.

As a quality control measure, each patient's DNA was also analyzed by SNP array using the Fluidigm SNPTrace panel (QC call rate > 90%). SNPtrace genotypes were compared to the GS data to confirm correct sample identification and assess sequencing quality. Samples passed quality control if the concordance between SNPtrace and GS was ≥ 90%.

The dataset included GS of 188 individuals, encompassing 75 participants along with their biological parents. In principle, each family was analyzed as a trio (except for two duo families consisting of the participant and their mother), composed of unaffected parents and an affected proband. In 12 families, affected siblings were included.

Sequencing data were processed through two complementary pipelines, reflecting the dual goals of clinical reporting and research discovery. For the clinical report, raw reads were analyzed using the Illumina DRAGEN Bio-IT Platform (1), which performs hardware-accelerated alignment to the GRCh38 reference genome, followed by variant calling of SNVs, indels, structural variants, and copy-number variants. To maintain consistency with clinical reporting standards, variants were subsequently lifted over from GRCh38 to the GRCh37 reference genome prior to clinical interpretation and reporting. Variants were interpreted by the clinical team under the ACMG guidelines (2) to generate diagnostic reports.

In parallel, a research-level analysis was performed to identify novel candidate genes. Sequencing reads were mapped to GRCh38 using the Burrows-Wheeler Aligner–Maximal Exact Matches (BWA-MEM) algorithm (3), and for SNV/indel analysis, trio-based variant calling was performed with Platypus (4). The resulting variants were annotated using ANNOVAR (5), incorporating both gene-level (RefSeq (6), UCSC genes (7), Swiss-Prot (8), OMIM (9)) and variant-level information. Variant-level annotations included predicted functional effects, population allele frequencies from gnomAD (v4.1) (10) and TOPMed (Freeze 8) (11), multiple in silico pathogenicity scores (REVEL (12), CADD (13), dbscSNV (14), AlphaMissense (15), SpliceAI (16)), and entries from disease and clinical variant databases such as OMIM and ClinVar (17). In addition, we utilized AI-MARRVEL (18) to prioritize candidate variants based on integrative gene- and variant-level pathogenicity predictions, incorporating genomic, transcriptomic and cross-species functional data.

For copy number variants (CNVs), five algorithms (BreakDancer (19), CNVnator (20), Delly (21), Lumpy (22), and manta (23) were incorporated. The resulting CNVs were annotated using ClassifyCNV (24), which integrates information on population frequency, overlap with protein-coding genes, ClinGen dosage sensitivity, and other genomic features relevant to pathogenicity assessment.

2) RNA Sequencing and Analysis

RNA sequencing (RNA-seq) was performed on peripheral blood from a subset of 63 cases to assess expression outliers and splicing abnormalities. Whole transcriptome sequencing (total RNA-seq) was performed in selected cases to evaluate expression outliers and splicing abnormalities. RNA libraries were prepared using the Illumina TruSeq Stranded Total RNA with Ribo-Zero Globin kit (Illumina, Cat# 20020612). Libraries were pooled in equimolar ratios (35 samples per pool) and sequenced on the NovaSeq 6000 platform with the S4 reagent kit (300 cycles) to generate 2×150 bp paired-end reads, targeting a minimum depth of 50 million read pairs (100 million total reads) per sample.

The RNA-seq analysis pipeline integrates open-source tools and internally developed protocols for data processing and quality control. Raw reads (FASTQs) were assessed using FastQC (v0.11.7) (25) and trimmed with TrimGalore (v0.6.5) (26) to remove 5′ adapters. Adapter-free reads were aligned to the GRCh37 reference genome using STAR (v2.7.3a) (27) to ensure compatibility with a previously established control dataset, and gene and isoform quantification was performed with RSEM (v1.3.3) (28). Duplicate reads were marked using Picard (v2.18.4) (29). Quality control metrics were generated with RNA-SeQC (v1.1.9) (30) and the internally developed quality control script (31). Samples were required to meet the following criteria to pass QC: >100 million total reads, >70% mapping rate, <50% duplication rate, <10% ribosomal RNA fraction, >95% strand specificity, and >10-fold mean per-base coverage.

Aberrant expression and splicing events were identified using DROP (v1.1.4) (32), applying the OUTRIDER and FRASER modules, respectively, on a batch of 60 samples. For expression outliers (OUTRIDER) we used the following cutoffs: adjusted p-value < 1, and fold-change less than 0.7 or greater than 1.3., while for splicing outliers (FRASER) we used only adjusted p-value < 1. All outlier events were visually inspected using Integrative Genomics Viewer (IGV) (33).

3) Exon trapping analysis

A genomic fragment encompassing the exon of interest along with approximately 300 base pairs of flanking intronic sequence on either side was amplified and cloned into the pET01 exon trap vector (plasmid #218972) to preserve native splicing context. Two constructs were generated: a wild-type (WT) sequence and a mutant (MT) sequence harboring the deep intronic variant DNMT3A:c.492+61C. Cloning was performed using the NEBuilder HiFi DNA Assembly Master Mix (New England Biolabs) according to the manufacturer’s instructions.

HEK293 cells were maintained under standard culture conditions and transiently transfected with either WT or MT exon trap constructs at a concentration of 250 ng/µL. Cells were harvested 24 hours post-transfection for downstream analysis.

Total RNA was extracted using a TRIzol-based method followed by purification with the Norgen RNA purification kit (Norgen Biotek), according to the manufacturer’s protocol. Purified RNA was subsequently reverse transcribed into complementary DNA (cDNA) using the qScript cDNA Synthesis Kit (Quanta Biosciences).

Amplification of cDNA was performed using exon trap–specific primers (forward: 5′-GATCGATCCGCTTCCTGCCCC-3′; reverse: 5′-CTGCCGGGCCACCTCCAGTGC-3′) and RepliQa DNA polymerase (Quantabio). PCR was carried out for 28 cycles with an annealing temperature of 67°C, an extension temperature of 68°C, and an extension time of 10 seconds.

PCR products were sequenced using Oxford Nanopore Technologies platform, generating approximately 5,000 reads per sample. Sequencing reads were aligned to the reference exon trap construct to identify splice junctions and transcript isoforms. Splicing outcomes were quantified by calculating the relative proportion of reads corresponding to each splice variant, enabling direct comparison between WT and MT constructs.

4. **Variant interpretations by ACMG guidelines and novel gene candidate search**

Clinical information for all patients was obtained from the REDCap database and Epic chart reviews, and often additional details were collected through direct communication with the treating physicians. Final clinical reports were delivered to patients by genetic counselors and physicians at the participating hospital.

1) Clinical Report (Known Genes, ACMG Guidelines)

Variants were first evaluated in the context of established disease-associated genes. Variants were classified as pathogenic, likely pathogenic, variants of uncertain significance (VUS), likely benign or benign according to the ACMG guidelines (2) and correlation with patient’s clinical phenotypes and family history. Findings including pathogenic, likely pathogenic or VUSs that fully or partially explained the clinical phenotype were returned to the treating physicians as part of the clinical report. In addition, pathogenic or likely pathogenic variants meeting ACMG recommended secondary finding (34) were returned to patients who selected to receive such findings, regardless of phenotype correlation.

2) Research-Level Novel Gene Candidate Search

For cases that remained unsolved or only partially explained by pathogenic or likely-pathogenic variants in known genes, research-level analyses were performed to identify novel candidate genes.

SNVs and indels were first prioritized based on population frequency (<1%), inheritance pattern (de novo, rare biallelic, or X-linked), and predicted deleteriousness from multiple algorithms (CADD >25; REVEL >0.7; AlphaMissense: likely pathogenic; SpliceAI >0.2). For copy-number variants (CNVs; large deletions/duplications), a CNV was considered valid if it was supported by at least two of the five callers and demonstrated >80% reciprocal overlap across algorithms to reduce false-positive calls. CNVs were prioritized if they were rare (MAF <1%), located within protein-coding genes, overlapped at least one exon, and arose de novo; for familial cases, shared variants among affected individuals with similar phenotypes were considered. Variants recurrently reported as benign in curated databases or peer-reviewed publications were excluded from further analysis.

Candidate SNVs/indels and CNVs were then evaluated through gene-level biological and phenotypic review. This review considered the known or predicted function of the affected gene, relevance to the patient’s organ systems or clinical features, evidence from model organisms or functional studies, prior human reports or related biological mechanisms supporting disease relevance, and RNA-seq evidence when available. RNA-seq data were reviewed for aberrant expression or splicing events that supported candidate gene or variant interpretation. Candidate CNVs were visually inspected using IGV and Samplot (35).

Candidate genes with convergent support from variant-level evidence, gene function, phenotype relevance, and available molecular data were nominated for further investigation and potential functional modeling, with final decisions determined through collaborative discussions between the clinical and genomic analysis teams. Consensus-prioritized candidate genes were submitted to GeneMatcher and queried in available rare disease resources, including GREGoR and an institutional clinical genetics laboratory, to identify additional individuals with overlapping phenotypes and candidate variants in the same genes.

**5, Acetylcholinesterase (*ACHE*) activity assay**

1) Generation of human *ACHE* expression vector

Human *ACHE* cDNA clone (NM_000665.5) was obtained from an institutional cDNA clone collection, and the mutagenesis was performed using Q5 site-directed mutagenesis kit (NEB, E0554S, USA). For the in vitro cell-based assay, these entry clones were shuttled into the pcDNA-DEST40 vector (ThermoFisher Scientific, 12274015, USA) using a Gateway-compatible LR Clonase reaction. These three constructs, ACHE-Reference, ACHE-p.Q5Afs32* and ACHE-p.R167G in pcDNA-DEST40 vector, were validated by whole-plasmid sequencing with Plasmidsaurus Inc. (USA).

2) In vitro acetylcholinesterase activity assay

Human embryonic kidney 293T (HEK293T) cells were maintained in the laboratory. HEK293T cells were cultured in DMEM medium containing 10% fetal bovine serum (GenDEPOT, F0900-050, USA) and penicillin and streptomycin under a humidified atmosphere containing 5% CO2. For human *ACHE* cDNA transfection, cells were plated at 1 × 10^6^ cells on a poly-D-lysine-coated 6-well plate. Transient transfection was performed with Xfect-transfection reagent (Takara Bio, 631318, Japan) according to the manufacturer’s instructions. After 24 hours, the culture media were replaced with serum-free DMEM media, and the cells were incubated for an additional 48 hours before the culture media collection. ACHE activity was determined using Acetylcholinesterase Fluorescent Activity Kit (Thermo Fisher Scientific, EIAACHEF, USA) according to the manufacturer’s instructions. Briefly, the culture medium of each sample was diluted 1/10 or 1/100 and mixed with the detection reagent and Acetylthiocholine iodide as the substrate. The fluorescence intensity was measured after 20 minutes of incubation at 510 nm using a GloMax® Discover microplate reader (Promega Instrument, USA).

3) Statistical analyses

The experiment was run in triplicate, including no-transfection and lacZ as negative control. The fluorescent activity for each sample was also recorded in 3 different wells, and then the average value recorded for that sample was plotted and analyzed by a two-tailed unpaired t-test in GraphPad Prism software (version 10.5.0). Data were presented as the mean ± SEM (e.g. 5693 ± 577.3) with significance levels indicated as follows: **p*<0.05, ***p*<0.01, ****p*<0.001.

**Supplementary References**

1.	Behera S, Catreux S, Rossi M, Truong S, Huang Z, Ruehle M, et al. Comprehensive genome analysis and variant detection at scale using DRAGEN. Nat Biotechnol. 2025;43(7):1177-91.

2.	Richards S, Aziz N, Bale S, Bick D, Das S, Gastier-Foster J, et al. Standards and guidelines for the interpretation of sequence variants: a joint consensus recommendation of the American College of Medical Genetics and Genomics and the Association for Molecular Pathology. Genet Med. 2015;17(5):405-24.

3.	Li H, Durbin R. Fast and accurate short read alignment with Burrows-Wheeler transform. Bioinformatics. 2009;25(14):1754-60.

4.	Rimmer A, Phan H, Mathieson I, Iqbal Z, Twigg SRF, Consortium WGS, et al. Integrating mapping-, assembly- and haplotype-based approaches for calling variants in clinical sequencing applications. Nat Genet. 2014;46(8):912-8.

5.	Wang K, Li M, Hakonarson H. ANNOVAR: functional annotation of genetic variants from high-throughput sequencing data. Nucleic Acids Res. 2010;38(16):e164.

6.	O'Leary NA, Wright MW, Brister JR, Ciufo S, Haddad D, McVeigh R, et al. Reference sequence (RefSeq) database at NCBI: current status, taxonomic expansion, and functional annotation. Nucleic Acids Res. 2016;44(D1):D733-45.

7.	Haeussler M, Zweig AS, Tyner C, Speir ML, Rosenbloom KR, Raney BJ, et al. The UCSC Genome Browser database: 2019 update. Nucleic Acids Res. 2019;47(D1):D853-D8.

8.	Boutet E, Lieberherr D, Tognolli M, Schneider M, Bairoch A. UniProtKB/Swiss-Prot. Methods Mol Biol. 2007;406:89-112.

9.	Amberger JS, Hamosh A. Searching Online Mendelian Inheritance in Man (OMIM): A Knowledgebase of Human Genes and Genetic Phenotypes. Curr Protoc Bioinformatics. 2017;58:1 2 1- 2 12.

10.	Chen S, Francioli LC, Goodrich JK, Collins RL, Kanai M, Wang Q, et al. A genomic mutational constraint map using variation in 76,156 human genomes. Nature. 2024;625(7993):92-100.

11.	Taliun D, Harris DN, Kessler MD, Carlson J, Szpiech ZA, Torres R, et al. Sequencing of 53,831 diverse genomes from the NHLBI TOPMed Program. Nature. 2021;590(7845):290-9.

12.	Ioannidis NM, Rothstein JH, Pejaver V, Middha S, McDonnell SK, Baheti S, et al. REVEL: An Ensemble Method for Predicting the Pathogenicity of Rare Missense Variants. Am J Hum Genet. 2016;99(4):877-85.

13.	Rentzsch P, Witten D, Cooper GM, Shendure J, Kircher M. CADD: predicting the deleteriousness of variants throughout the human genome. Nucleic Acids Res. 2019;47(D1):D886-D94.

14.	Jian X, Boerwinkle E, Liu X. In silico prediction of splice-altering single nucleotide variants in the human genome. Nucleic Acids Res. 2014;42(22):13534-44.

15.	Cheng J, Novati G, Pan J, Bycroft C, Zemgulyte A, Applebaum T, et al. Accurate proteome-wide missense variant effect prediction with AlphaMissense. Science. 2023;381(6664):eadg7492.

16.	Jaganathan K, Kyriazopoulou Panagiotopoulou S, McRae JF, Darbandi SF, Knowles D, Li YI, et al. Predicting Splicing from Primary Sequence with Deep Learning. Cell. 2019;176(3):535-48 e24.

17.	Landrum MJ, Lee JM, Benson M, Brown GR, Chao C, Chitipiralla S, et al. ClinVar: improving access to variant interpretations and supporting evidence. Nucleic Acids Res. 2018;46(D1):D1062-D7.

18.	Mao D, Liu C, Wang L, Ai-Ouran R, Deisseroth C, Pasupuleti S, et al. AI-MARRVEL - A Knowledge-Driven AI System for Diagnosing Mendelian Disorders. NEJM AI. 2024;1(5).

19.	Chen K, Wallis JW, McLellan MD, Larson DE, Kalicki JM, Pohl CS, et al. BreakDancer: an algorithm for high-resolution mapping of genomic structural variation. Nat Methods. 2009;6(9):677-81.

20.	Abyzov A, Urban AE, Snyder M, Gerstein M. CNVnator: an approach to discover, genotype, and characterize typical and atypical CNVs from family and population genome sequencing. Genome Res. 2011;21(6):974-84.

21.	Rausch T, Zichner T, Schlattl A, Stutz AM, Benes V, Korbel JO. DELLY: structural variant discovery by integrated paired-end and split-read analysis. Bioinformatics. 2012;28(18):i333-i9.

22.	Layer RM, Chiang C, Quinlan AR, Hall IM. LUMPY: a probabilistic framework for structural variant discovery. Genome Biol. 2014;15(6):R84.

23.	Chen X, Schulz-Trieglaff O, Shaw R, Barnes B, Schlesinger F, Kallberg M, et al. Manta: rapid detection of structural variants and indels for germline and cancer sequencing applications. Bioinformatics. 2016;32(8):1220-2.

24.	Gurbich TA, Ilinsky VV. ClassifyCNV: a tool for clinical annotation of copy-number variants. Sci Rep. 2020;10(1):20375.

25.	Andrews S. FastQC: A Quality Control Tool for High Throughput Sequence Data [Online] 2010 [Available from: http://www.bioinformatics.babraham.ac.uk/projects/fastqc/.

26.	Krueger F. FelixKrueger/TrimGalore: v0.6.7. Zenodo. 2021 [

27.	Dobin A, Davis CA, Schlesinger F, Drenkow J, Zaleski C, Jha S, et al. STAR: ultrafast universal RNA-seq aligner. Bioinformatics. 2013;29(1):15-21.

28.	Li B, Dewey CN. RSEM: accurate transcript quantification from RNA-Seq data with or without a reference genome. BMC Bioinformatics. 2011;12:323.

29.	Institute B. "Picard Toolkit." GitHub Repository.

30.	DeLuca DS, Levin JZ, Sivachenko A, Fennell T, Nazaire MD, Williams C, et al. RNA-SeQC: RNA-seq metrics for quality control and process optimization. Bioinformatics. 2012;28(11):1530-2.

31.	Center BCoMHGS. ERCCQC 2019 [Available from: <https://github.com/BCM-HGSC/ERCCQC>.

32.	Yépez VA, Mertes C, Mueller MF, Andrade DS, Wachutka L, Frésard L, et al. Detection of aberrant gene expression events in RNA sequencing data. 2019.

33.	Robinson JT, Thorvaldsdottir H, Winckler W, Guttman M, Lander ES, Getz G, et al. Integrative genomics viewer. Nat Biotechnol. 2011;29(1):24-6.

34.	Miller DT, Lee K, Abul-Husn NS, Amendola LM, Brothers K, Chung WK, et al. ACMG SF v3.2 list for reporting of secondary findings in clinical exome and genome sequencing: A policy statement of the American College of Medical Genetics and Genomics (ACMG). Genet Med. 2023;25(8):100866.

35.	Belyeu JR, Chowdhury M, Brown J, Pedersen BS, Cormier MJ, Quinlan AR, et al. Samplot: a platform for structural variant visual validation and automated filtering. Genome Biol. 2021;22(1):161.
