## Supplementary figures and images for "Integrated Clinical-Genomic Reanalysis Identifies Diagnostic Gaps and Candidate Genes in a Pediatric Rare Disease Cohort"

### Supplementary Figure 1

**Supplementary Figure 1. Interval to reanalysis by outcome group**

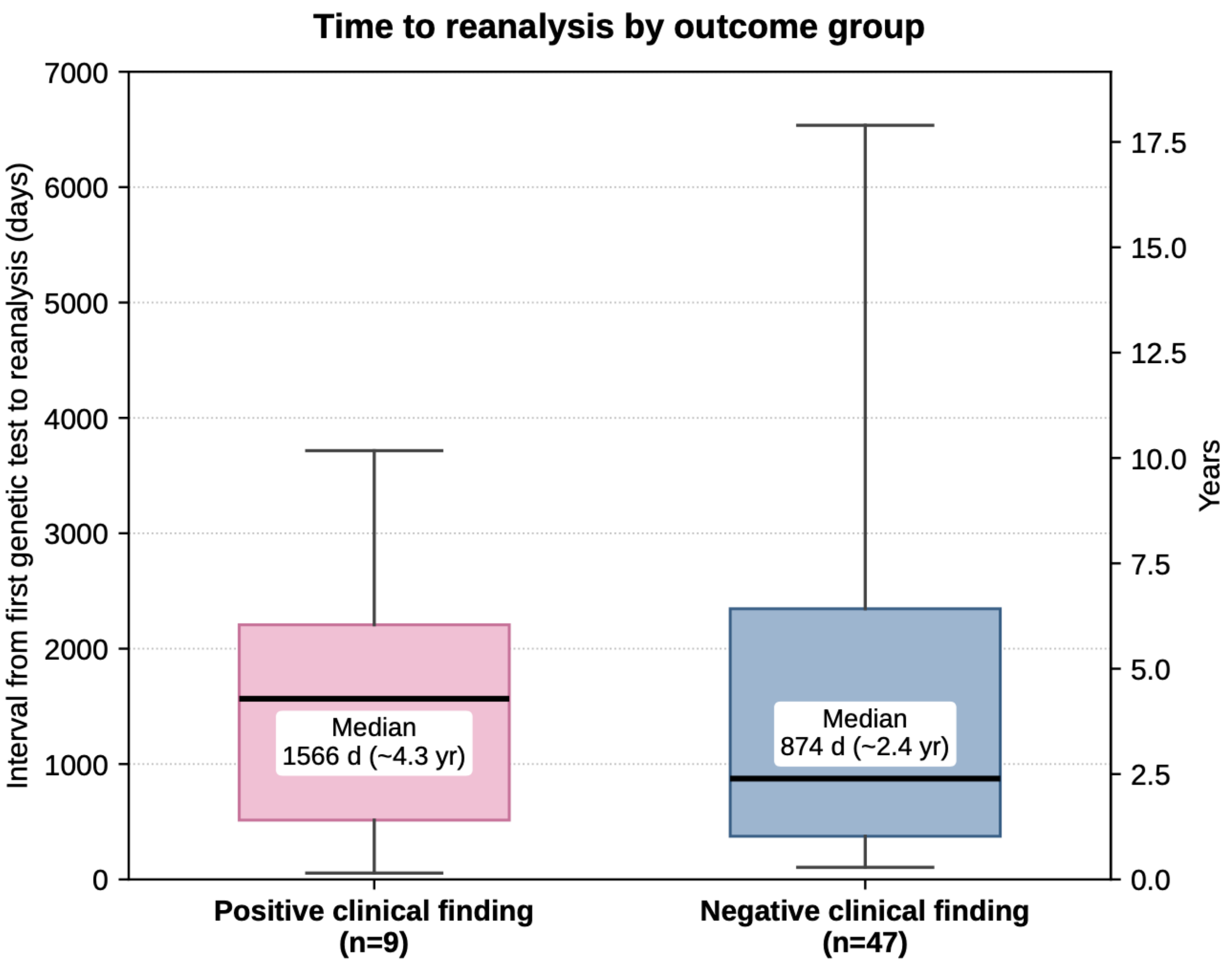


Two-sided Mann–Whitney $P = 0.41$.
