## Supplementary Table 2 for "Integrated Clinical-Genomic Reanalysis Identifies Diagnostic Gaps and Candidate Genes in a Pediatric Rare Disease Cohort"

| **Category / Feature** | **Case in This Study (Presumed Functional ACHE Deficiency)** | **Cholinergic Crisis**  **(Acute Excess)** | **Congenital Myasthenic Syndrome (COLQ-related)** |
| --- | --- | --- | --- |
| **Proposed Pathophysiologic Mechanism** | Chronic partial enzymatic acetylcholinesterase deficiency due to biallelic *ACHE* variants | Acetylcholine excess by acute acetylcholinesterase inhibition | Impaired synaptic anchoring of acetylcholinesterase due to COLQ deficiency |
| **Temporal Pattern** | Chronic, multisystem | Acute, rapidly progressive | Chronic, neuromuscular |
| **Seizure** | Yes | Possible | Rare |
| **Drooling / Salivation** | Yes | Prominent | Occasional |
| **Miosis** | Yes | Yes | No |
| **Ptosis / Ophthalmoplegia** | No | No | Yes |
| **Generalized Hypotonia / Weakness** | Yes | Yes | Yes (fatigable) |
| **Myoclonus / Fasciculations** | Yes | Common | Not typical |
| **Autonomic GI Dysfunction** | Yes (constipation-predominant, dysphagia) | Yes (diarrhea) | Occasional |
| **Abnormal Reaction to Anesthesia** | Yes (delayed recovery) | Possible (enhanced sensitivity) | Reported (perioperative vulnerability) |
| **Developmental Delay / Cognitive Impairment** | Yes | No | Usually, normal cognition |
| **Scoliosis** | Yes | No | Occasional |
| **Overall Clinical Course** | Chronic, multisystem, non-lethal | Acute, life-threatening | Chronic, often treatable |

**Supplementary Table 2. Comparison of clinical features in the case in this study and representative acetylcholinesterase-related or cholinergic disorders**
